# A Phase 2, Randomized, Double-blind, Placebo-controlled Study of oral RP7214, a DHODH inhibitor, in Patients with Symptomatic Mild SARS-CoV-2 Infection

**DOI:** 10.1101/2023.02.08.23285565

**Authors:** Ajit Nair, Prajak Barde, Kasi V Routhu, Swaroop Vakkalanka, RP7214-2101 Study Group

## Abstract

**Introduction:** COVID-19 pandemic due to severe acute respiratory syndrome coronavirus 2 (SARS-CoV-2) is of immense global public health concern. RP7214, a novel, potent, oral, inhibitor of DHODH, has shown preclinical evidence in inhibiting viral replication and lung inflammation.

**Methods:** This was a randomized, double-blind, placebo-controlled phase 2 study in patients with symptomatic mild SARS-CoV-2 infection, having at least one high-risk feature (e.g., hypertension, diabetes mellitus) for developing severe Covid-19 infection. The patients received RP7214 (400 mg BID) or a placebo for 14 days in a blinded fashion and were followed up to 30 days. Patients also received supportive therapy (e.g., antipyretics and antitussives for symptomatic relief) at the discretion of the investigator. The endpoints were Covid 19 related hospitalization rate by Day 15, SARS-CoV-2 viral load and clearance on Days 3,7 and 15, clinical symptoms improvement by Day 15, safety, and the immuno-modulatory effect of RP7214.

**Results:** A total of 163 patients were treated in the study; 82 received RP7214 and 81 received placebo. Of the total patients, 44.2% had received Covid-19 vaccine prior to the study. The symptom onset was ≤ 3 days in 22.1%. None of the patients in the study required hospitalization. There was no difference in the mean change of viral load between RP7214 and placebo. In the subgroup analysis, in patients having symptom onset of ≤ 3 days, RP7214 significantly reduced viral load on Days 3 and 7, respectively. Similarly, in non-vaccinated patients with symptom onset of ≤ 3 days, RP7214 significantly reduced viral load on Day 3. Overall, there was a trend towards better viral load reduction in RP7214-treated patients with a baseline viral load of 5 log units or higher. For all other endpoints, there was no difference between RP7214 and placebo. Majority of the reported AEs were mild and not related either to study treatment.

**Conclusions:** RP7214 at 400 mg BID dose level showed a statistically significant reduction in viral load at an early stage of the disease and in non-vaccinated patients. There was a trend towards better viral load reduction in RP7214-treated patients with a baseline viral load of 5 log units or higher. RP7214 showed a favorable safety profile. Further development of RP7214 in Covid 19 in a mild symptomatic population with co-morbidities and treated at an early stage of disease may show benefit.

## Introduction

The emergence of severe acute respiratory syndrome coronavirus 2 (SARS-CoV-2) triggered an ongoing global pandemic of COVID-19.^[1]^ As of 23^rd^ December 2022, there have been 651,918,402 confirmed cases of COVID-19, including 6,656,601 deaths as reported to WHO.^[2]^

An early step in the COVID-19 infection is the interaction of the spike protein on the surface of the virus with angiotensin-converting enzyme 2 (ACE2) receptors on the surface of the host cell. After entry into the host cell, the virus uses components of the host cell to replicate and secrete viral particles and disrupt RNA handling and protein translation. ^[3, 4]^ The rapid and uncontrolled viral replication of SARS-CoV2 escapes the host’s innate immune activation during its initial steps. As a result, the increase in pro-inflammatory responses and immune cellular infiltration in the lungs leads to tissue damage and contributes to the clinical manifestation of SARS-CoV-2.^[5]^ This inflammation can result in extreme permeability of the vasculature, multi-organ failure, acute respiratory distress syndrome (ARDS), and death.^[6]^ Thus, in the early stages of infection, the primary target of drugs is to block or reduce viral replication and control the inflammatory reactions in the lung. A therapeutic agent that can inhibit SARS-CoV-2 replication and attenuate lung inflammation can be beneficial to treat COVID-19 infection.

Dihydroorotate dehydrogenase (DHODH) is a ubiquitous enzyme essential for the *de novo* production of pyrimidines. It catalyzes the conversion of dihydroorotate to orotate and is the only enzyme capable of performing this conversion. Inhibitors of DHODH function by blocking the conversion of dihydroorotate to orotate leads to the rapid depletion of UMP, UDP, and UTP which are the building blocks of RNA and DNA biosynthesis. Rapidly proliferating cells, such as lymphocytes and viruses including the SARS-CoV-2, depend on *de novo* pyrimidine biosynthesis to support their replication. This indicates that inhibition of DHODH, a rate-limiting enzyme can become a key step in the treatment of COVID-19 infection.^[7]^

RP7214 is a novel, potent, orally available inhibitor of the DHODH enzyme. It blocks the conversion of L-DHO to orotic acid, with an IC_50_ of 7.8nM in THP-1 cells. It also inhibits PHA-induced CD4 + cell proliferation with IC_50_ of 2.45 nM in human whole blood and 0.60 nM in human PBMC.^[3]^

RP7214 showed anti-viral activity in Vero-E6 infected SARS-CoV-2 with an IC_50_ of 1.51 μM and 1.22 μM for the E gene and N gene, respectively. It showed more than 90% inhibition at 5 μM without affecting the viability of Vero-E6 cells.^[8]^

At a dose of 30 mg/kg, oral RP7214 reduced LPS-induced neutrophil infiltration into the bronchoalveolar lavage fluid in rats at 24 h with an efficacy similar to dexamethasone. In Balb/c mice, RP7214 showed a dose-dependent reduction in plasma levels of the pro-inflammatory cytokines and IL-17, following a 2 hr stimulation with 20 mg/kg Concanavalin A. These studies suggest the anti-inflammatory activity of RP7214.^[9]^

A phase 1, single Ascending study (SAD) study conducted in healthy subjects at 100, 200, and 400 mg QD doses and multiple Ascending study (MAD) study conducted at 200 and 400 mg BID doses given for 7 days demonstrated that RP7214 was well tolerated and with no major safety findings at any of the dose levels. Pharmacokinetics (PK) at single doses of 100, 200, and 400 mg showed rapid absorption of RP7214 with the maximum peak in about 2-3 hours followed by quick elimination with a half-life ranging from 5-7 hours. Steady-state concentrations were achieved in about 5-6 days of dosing.^[9]^

The rationale for dose selection for this study was based on achieving a target concentration in the lung that exceeds the IC_50_ of RP7214 for inhibiting both the E & the N gene in *in-vitro* studies. Assuming an average lung to the serum concentration ratio of ∼0.1 in the animal study, steady-state plasma concentrations of 78.40 μM achieved with 400 mg BID dose were considered sufficient to show efficacy in patients with mild Covid-19 infection.

The favourable data derived from the preclinical experiments and phase 1 studies show that RP7214 has the potential in treating Covid 19. Hence, this phase 2 study was undertaken to evaluate the efficacy and safety of RP7214 in patients with symptomatic mild SARS-CoV-2 infection.

## Materials and Methods

### Study design

This was a phase 2, double-blind, placebo-controlled study conducted between September 2021 to March 2022. The primary objective was to evaluate the effect of RP7214 on Covid-19-related hospitalization as compared to a placebo. The secondary objectives were to characterize the effect of RP7214 on SARS-CoV-2 viral load and clearance, its effect on clinical symptoms, safety, and immuno-modulatory effect.

The total duration of the subject participation was 30 (± 2) days including a screening period of -2 to Day 1, a treatment period of 14 days, and a follow-up period extending up to Day 30. All patients were in home isolation or quarantined for the duration of the study and hence all study assessments/visits except screening and End of Study (EOS) were done virtually using video calls or telephone calls.

### Ethics

The study was conducted in compliance with the clinical trial protocol approved by the Drug Controller General of India (DCGI) and the Institutional Ethics Committee(s) (IEC). The study complied with the International Council for Harmonisation (ICH) Good Clinical Practice (GCP) guidelines and the current revision of the Declaration of Helsinki, ‘Indian GCP guidelines’; and ‘New Drugs and Clinical Trials Rules 2019’; and ‘Ethical Guidelines for Biomedical Research on Human Subjects’ issued by the Indian Council of Medical Research in 2017. The study was registered in the Clinical Trial Registry of India (CTRI) before the start of enrolment. All subjects provided written informed consent prior to screening.

The protocol, informed consent form(s), and all patient materials were approved by all seventeen Institutional/Independent Ethics Committees (EC). The Institutional Human Ethics Committee of Panimalar Medical college hospital and research Institute, Chennai, India gave first ethics approval for this work which was received on 08 Oct 2021 to conduct study at Panimalar Medical college hospital and research Institute.

### Study participants

Male and female patients aged ≥ 18 years with mild Covid-19 infection having ≥ 1 symptom (fever, cough, sore throat, malaise, headache, muscle pain, nausea, vomiting, diarrhea, loss of taste and/or smell) without any shortness of breath or hypoxia (Respiratory rate should be < 24/min) and SpO_2_ ≥ 94% on room air were included in the study. Patients with a laboratory-confirmed Covid-19 infection by Reverse Transcription Polymerase Chain Reaction (RT-PCR) in the nasopharyngeal sample (within 72 hours prior to randomization) and with at least one pre-existing high-risk feature (e.g., age > 60 years, hypertension, diabetes mellitus, chronic lung disease, chronic kidney disease, liver disease, cerebrovascular disease, obesity (body mass index (BMI) > 30.0 kg/m^2^), cancer) for developing severe Covid-19 illness, were included in the study.

Patients with asymptomatic Covid-19 infection, onset of Covid-19 symptoms > 5 days at the time of randomization, moderate to severe Covid-19 infection, current use of other DHODH inhibitors including teriflunomide or leflunomide, or on or immediately requiring Covid-19 directed treatment such as antivirals (e.g., remdesivir, favipiravir), immunomodulatory agents (e.g., tocilizumab, itolizumab, baricitinib or JAK inhibitors), convalescent plasma, oral/intravenous steroids, or monoclonal antibodies at the time of screening were excluded from the study. Also, patients who were severely immunocompromised and had autoimmune diseases, or any bleeding disorders were excluded from the study.

### Study treatment

Eligible patients were randomized to one of the two groups to receive either RP7214 [400 mg BID] or a placebo. RP7214/Placebo tablets were self-administered orally twice a day for 14 days. Food and fluid intake other than water was restricted one hour before and one hour after the administration of the drug. Patients in both groups received supportive therapy for Covid-19 related signs/symptoms, including antipyretics (e.g., paracetamol) and anti-tussive (anti-tussive formulations were corticosteroid free), or inhaled budesonide for cough, nutritional supplements at the discretion of the treating physician and according to guidelines for the management of Covid-19 patients by DGHS, MoHFW, Government of India.^[10]^

### Restriction

The study restricted the use of antivirals (lopinavir/ritonavir combination, remdesivir, favipiravir), other DHODH inhibitors (teriflunomide and leflunomide), oral/intravenous steroids, ivermectin, monoclonal antibodies, immunomodulatory agents (chloroquine, hydroxychloroquine, tocilizumab, sarilumab, itolizumab, baricitinib, or JAK inhibitors), convalescent Covid-19 plasma and any other investigational agent. If prohibited medications were required as rescue treatments, patients were to be immediately discontinued from the study and were to be treated as per the institutional protocol. Rescue treatment will not be provided as a part of the study.

### Randomization & blinding

Randomization to either RP7214 or placebo was performed using an Interactive Web Response System (IWRS). The randomization schedule was generated using SAS® (SAS Institute Inc., USA) by the statistician. The treatment allocation codes corresponding to each patient randomized in the trial were maintained centrally. There were no medical emergencies, during the trial hence there was no request for code breaks by the investigator.

### Efficacy assessment

Efficacy assessments captured the proportion of patients requiring Covid-19-related hospitalization by Day 15, change from baseline in SARS-CoV-2 viral load on Days 3, 7, and 15, time to symptom resolution, proportion of patients demonstrating symptom resolution on Days 3, 7, and 15 using a standard “Symptoms Evolution of Covid-19 (SE-C19) Scale”;^[11-14]^ time to symptom improvement on Days 3, 7, and 15.

### Other assessments

Other assessment includes analysis of viral load clearance (proportion of patients who were negative on Days 3, 7, and 15 and time to viral load negativity), change from baseline in total symptom score on Days 3, 7, and 15 compared to placebo, the correlation between change from baseline in viral load and symptom score, assessment of change from baseline in SARS-CoV-2 viral RNA amongst patients who had onset of symptom with ≤ 3 days and in patients with onset of > 3 days.

### Safety assessment

All patients were followed for safety from the enrolment to EOS. Safety parameters included AEs, related AEs, AEs leading to discontinuation, serious AEs, and death. Parameters also included evaluation of clinical laboratory tests, vital signs, oxygen saturation (SpO_2_), physical examination, and ECG changes. Common Terminology Criteria for Adverse Events (CTCAE) Version 5.0 was used for the assessment of severity. Laboratory tests were performed on Days 1, 3, 7, and 15. Clinical status and treatment compliance was recorded till Day 15. Adverse Event (AE)/ Serious Adverse Event (SAE) assessment was done daily till Day 15 and then at the EOS visit.

### Biomarker assessment

Blood samples were collected from all patients for analysis of disease-related markers on Days 1, 3, 7, and 15, and were compared to the baseline. The disease-specific inflammatory markers included ferritin, C-reactive protein (CRP), D-dimer, LDH, and IL-6.

### Statistical analysis

All patients who received at least a single dose of the study medication were included in the safety population. All patients who were randomized and had received at least a single dose of the study medication and provided at least one post-baseline measure for the relevant endpoint were part of the modified intent-to-treat (mITT) population. mITT population was further divided into 4 different subsets. These subsets were defined as the mITT1, the population which included patients who had baseline SARS-CoV-2 viral RNA titer > 350 genome equivalents/mL (Log value: > 2.54), mITT2 population which included patients who had baseline SARS-CoV-2 viral RNA titer ≥ 1000000 genome equivalents/mL (Log value ≥ 5), mITT3 population which included patients who had baseline SARS-CoV-2 viral RNA titer < 1000000 to > 350 genome equivalents/mL (Log value between > 2.54 and 5). All mITT patients without major protocol deviations were considered under the per-protocol population.

The sample size calculation assumed a relative risk of reduction of Covid-19-related hospitalization of about 83% with RP7214 over placebo. Based on this, a total of 204 enrolled patients (102 patients per group) would have provided 80% power to detect a significant treatment difference at the two-sided significance level of 0.05.

Patients with missing SARS-CoV-2 viral RNA at a visit(s) post baseline were imputed based on the available non-missing SARS-CoV-2 viral RNA data. Patients with missing clinical symptom score data were imputed using either baseline observation carried forward (BOCF) or last observation carried forward (LOCF) approach. For patients with missing baseline or post-baseline laboratory biomarkers, no imputations were performed.

SARS-CoV-2 viral RNA measured in genome equivalents/mL were converted to log values. The change in SARS CoV-2 viral load from baseline to Day 3, 7, and 15 were compared between two treatment groups by repeated measure ANCOVA. For the subgroup analysis, two-sample t-test(t) or Mann-Whitney U-test(U) was used depending on the normality of the data. Adjusted least square mean and the standard error values along with the p-value and the 95% confidence interval of the difference between treatment arms were reported. The median time to symptom improvement/resolution was computed with 95% CI using the Kaplan-Meier method by treatment groups. A comparison of the distribution of improvement/resolution time was done through a log-rank test. The frequency and percentage of patients who demonstrate symptom improvement/resolution were calculated by using the Clopper-Pearson method. The difference in proportion of patients who were negative on Days 3, 7, and 15 between the RP7214 and placebo groups was computed along with 95% CI using Mietinen-Nurminen method.

## Results

### Subjects disposition and demographics

The study planned to enroll a total of 204 patients, but due to waning of the pandemic, only 167 patients were screened. Out of the 163 patients, 82 received RP7214 and 81 received placebo. A total of 160 patients (98.2%) completed the study and 3 patients discontinued the study. Most of the enrolled patients were males (70.6%). The overall average age, height, and weight were 4.82 years, 1.6 m, and 70.2 kg, respectively, and comparable in both groups (**Table 1**).

**Table 1:**
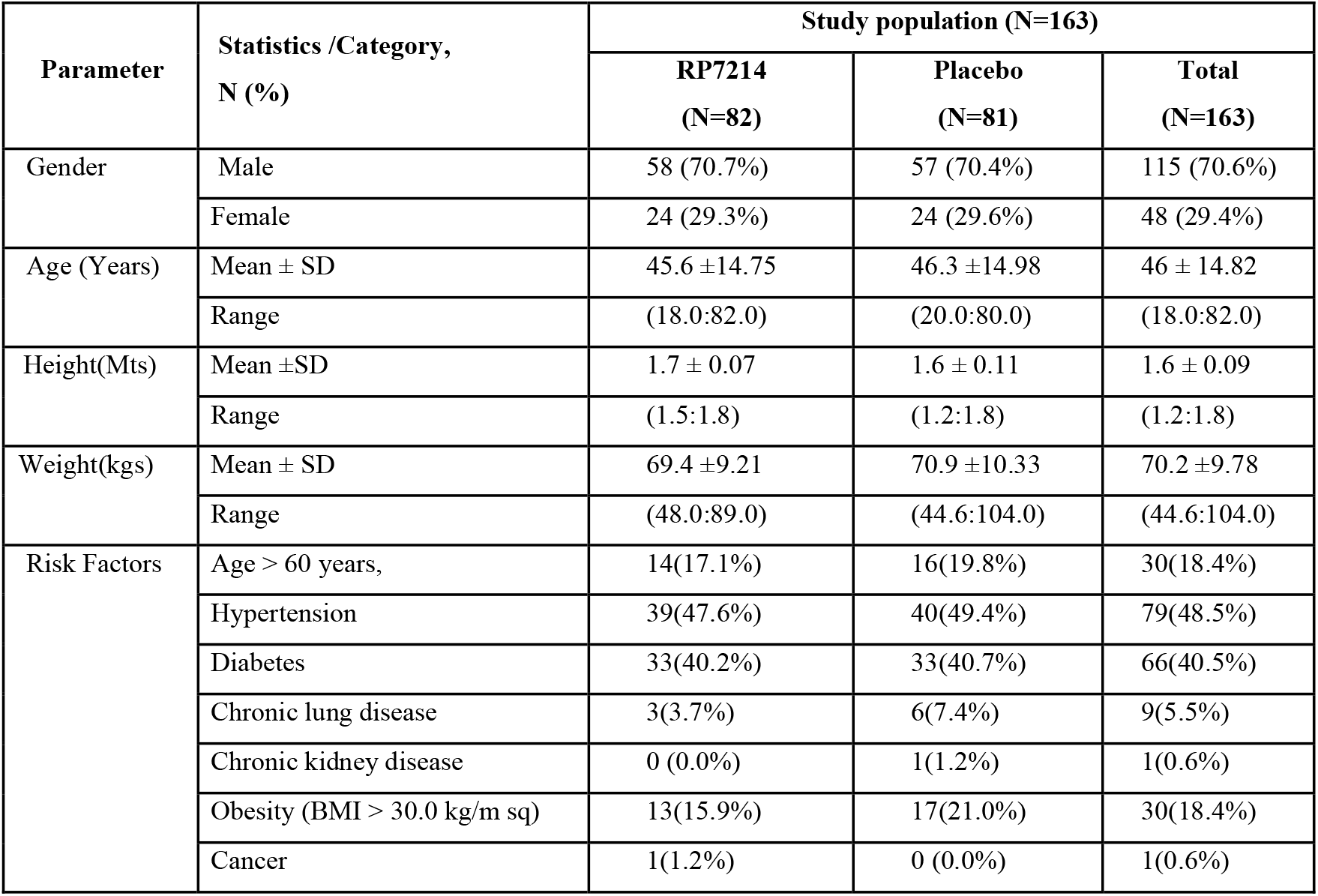
Summary of Subject Demography and Risk Factors (N=163)

| Parameter | Statistics /Category,<br>N (%) | Study population (N=163) |  |  |
| --- | --- | --- | --- | --- |
|  |  | RP7214<br>(N=82) | Placebo<br>(N=81) | Total<br>(N=163) |
| Gender | Male | 58 (70.7%) | 57 (70.4%) | 115 (70.6%) |
|  | Female | 24 (29.3%) | 24 (29.6%) | 48 (29.4%) |
| Age (Years) | Mean ± SD | 45.6 ± 14.75 | 46.3 ± 14.98 | 46 ± 14.82 |
|  | Range | (18.0:82.0) | (20.0:80.0) | (18.0:82.0) |
| Height(Mts) | Mean ±SD | 1.7 ± 0.07 | 1.6 ± 0.11 | 1.6 ± 0.09 |
|  | Range | (1.5:1.8) | (1.2:1.8) | (1.2:1.8) |
| Weight(kgs) | Mean ± SD | 69.4 ± 9.21 | 70.9 ± 10.33 | 70.2 ± 9.78 |
|  | Range | (48.0:89.0) | (44.6:104.0) | (44.6:104.0) |
| Risk Factors | Age > 60 years, | 14(17.1%) | 16(19.8%) | 30(18.4%) |
|  | Hypertension | 39(47.6%) | 40(49.4%) | 79(48.5%) |
|  | Diabetes | 33(40.2%) | 33(40.7%) | 66(40.5%) |
|  | Chronic lung disease | 3(3.7%) | 6(7.4%) | 9(5.5%) |
|  | Chronic kidney disease | 0 (0.0%) | 1(1.2%) | 1(0.6%) |
|  | Obesity (BMI > 30.0 kg/m sq) | 13(15.9%) | 17(21.0%) | 30(18.4%) |
|  | Cancer | 1(1.2%) | 0 (0.0%) | 1(0.6%) |

### Baseline characteristics

Out of the enrolled patients, 48.5% were hypertensive, 40.5% were diabetic, 18.4% were over 60 years of age, 18.4% were obese, 5.5% had chronic lung disease, 1 patient each had chronic kidney disease and breast cancer. The distribution of patients with the risk factors was similar in both treatment groups (**Table 2**).

**Table 2:** Summary of Baseline Characteristics.

| Baseline Character, n (%) | Study population (N=163) |  |  |
| --- | --- | --- | --- |
|  | RP7214<br>(N=82) | Placebo<br>(N=81) | Total<br>(N=163) |
| **Covid-19 symptom** |  |  |  |
| • Duration of symptom onset ≤ 3 days | 20(24.4%) | 16(19.8%) | 36(22.1%) |
| • Duration of symptom onset $>$ 3 days | 62(75.6%) | 65(80.2%) | 127(77.9%) |
| Patients with prior Covid-19 infection | 1(1.2%) | 3(3.7%) | 4(2.5%) |
| • Severity of infection: mild | 0(0.0%) | 2(2.5%) | 2(1.2%) |
| • Severity of infection: moderate | 1(1.2%) | 0(0.0%) | 1(0.6%) |
| Patients with a travel history | 8(9.8%) | 4(4.9%) | 12(7.4%) |
| No. patients who had contact with any Covid-19 patient | 17(20.7%) | 15(18.5%) | 32(19.6%) |
| Receipt of Covid-19 vaccine prior to the study | 40(48.8%) | 32(39.5%) | 72(44.2%) |
| • No. of doses received: One | 7(8.5%) | 3(3.7%) | 10(6.1%) |
| • No. of doses received: Two | 33(40.2%) | 29(35.8%) | 62(38.0%) |
| **Clinical symptoms at baseline** |  |  |  |
| Number of symptoms (N) | 82 | 81 | - |
| Mean ± SD | 5.00 ± 1.81 | 5.17 ± 2.14 | - |
| Range | (2.0:9.0) | (3.0:15.0) | - |
| Patients who reported a loss of taste, n (%) | 38(46.34%) | 40(49.38%) | - |
| Patients who reported a loss of smell, n (%) | 44(53.66%) | 32(39.51%) | - |
| Patients who reported fatigue, n (%) | 34(41.46%) | 32(39.51%) | - |

The duration of symptom onset was ≤ 3 days in 22.1% of patients while 77.9% of patients had symptom onset >3 days. Only 4 patients (2.5%) had a history of prior Covid-19 infection. None of the patients had prior Covid-19 infection more than once. Of the total patients, 44.2% had received a Covid-19 vaccine prior to the study. The average number of clinical symptoms at the baseline in both groups was comparable. The clinical symptoms at baseline, such as loss of taste, smell, and fatigue, were similar between the treatment groups (**Table 2**).

A total of 163 patients (RP7214: 82, and Placebo: 81) were enrolled in the different mITT and safety populations. The subgroup population included 77 patients (47.2%) in the mITT1 population (RP7214: 38, and Placebo: 39]; 43 patients (26.4%) in the mITT2 population (RP7214: 21 and Placebo: 22); 34 patients (20.9%) in the mITT3 population (RP7214: 17 and Placebo: 17). A total of 128 patients (78.5%) were randomized as PP population (RP7214: 60 and Placebo: 68).

### Efficacy

#### Covid-19 related hospitalization

None of the patients in either the RP7214 or the placebo groups showed worsening of disease that required hospitalization by Day 15.

#### Change in SARS-CoV-2 viral load

Out of 163 enrolled patients, 47.2% of patients had a baseline viral load of >2.4 log units and were considered as mITT1 population for analysis.

There was no statistically significant difference in the mean change of viral load from baseline on Days 3, 7, and 15 between RP7214 and the placebo group in the PP and overall mITT populations (**Figure 1**).

**Figure 1:**
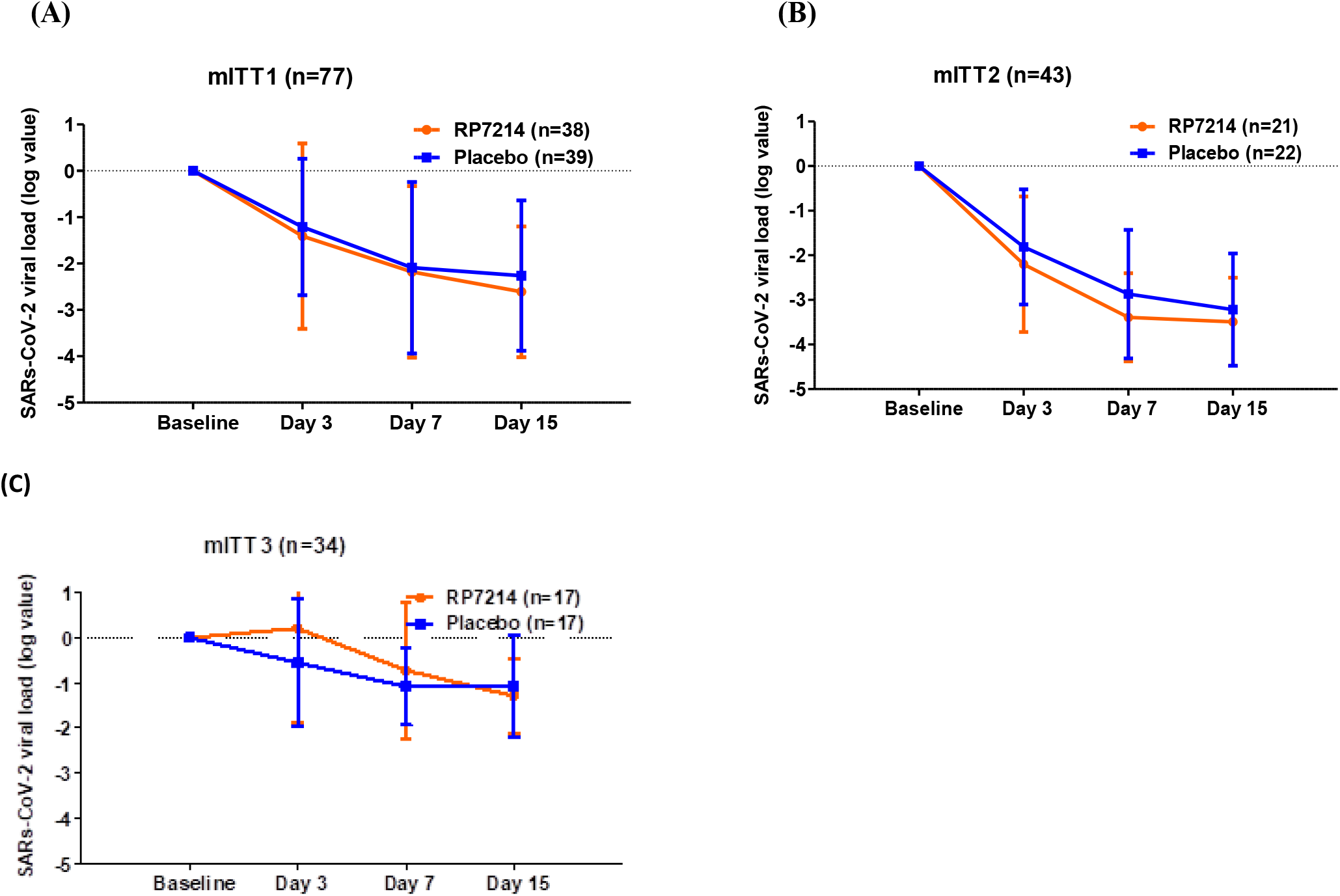
Change from baseline in SARS-CoV-2 viral titer for mITT1 population (A), mITT2 population (B), and mITT3 populations (C) Data are presented as means ±SDs. The change in SARS-CoV-2 viral load from baseline to Days 3, 7, and 15 was analyzed using the repeated measure ANCOVA Model.

**Figure 2:**
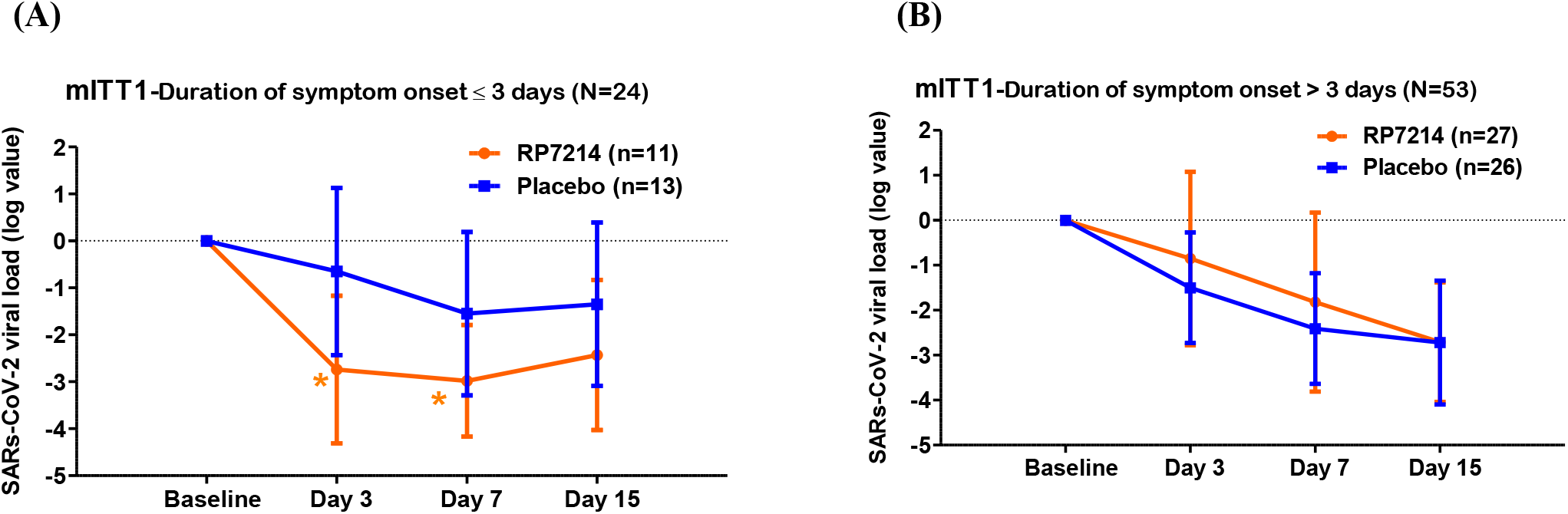
Change from baseline in SARS-CoV-2 viral titer for the mITT1 population with a duration of symptom onset ≤ 3 days (A) and mITT1 population with a duration of symptom onset > 3 days (B) Data are presented as means ±SDs. The change in SARS-CoV-2 viral load from baseline to Days 3, 7, and 15 was analyzed using the repeated measure ANCOVA Model. * p-value compares LS means of change from baseline between active and placebo arms.

No statistically significant difference in the mean change of viral load from baseline on Days 3, 7, and 15 was seen in the mITT1 vaccinated and non-vaccinated populations. (**Figure 3)**

**Figure 3:**
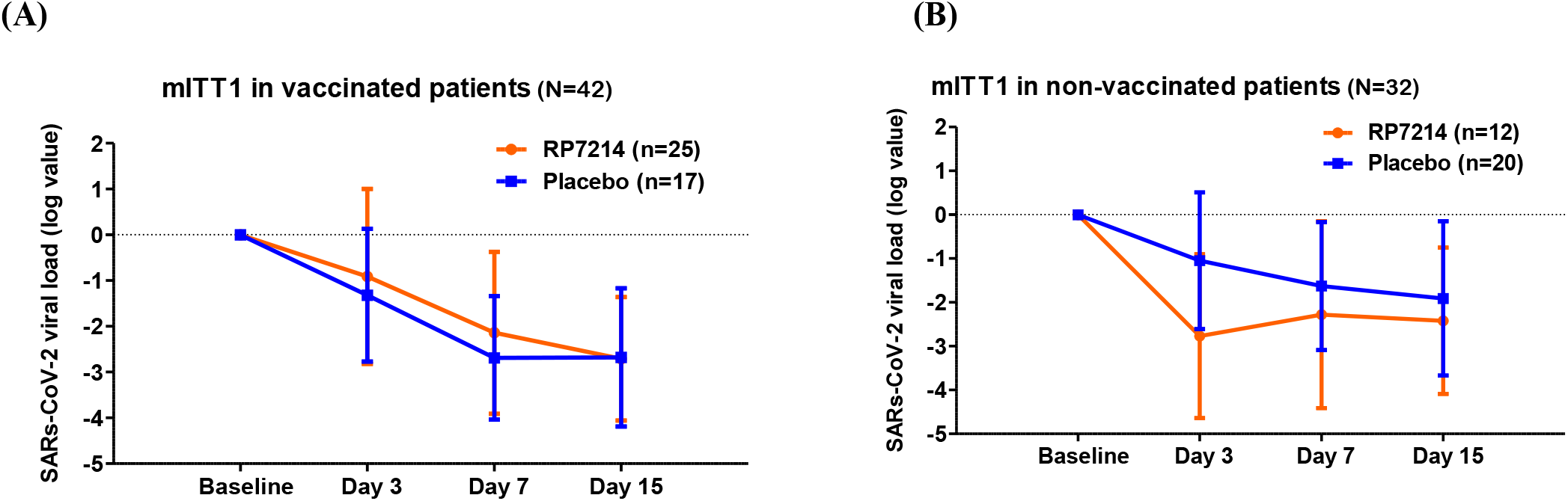
Change from baseline in SARS-CoV-2 viral titer for mITT1 vaccinated (A) and mITT1 non-vaccinated (B) Data are presented as means ±SDs. The change in SARS-CoV-2 viral load from baseline to Days 3, 7, and 15 was analyzed using the repeated measure ANCOVA Model.

When the change in viral load was assessed based on the duration of symptom onset (≤ 3 or > 3 days), in the mITT1 patients having symptom onset of ≤ 3 days, RP7214 significantly reduced viral load by a mean of an additional -2.09 (p= 0.0136), -1.43 (p= 0.0265) log genomic equivalents on Days 3 and 7, respectively (**Table 3, Figures 2**).

**Table 3:** Change from baseline in SARS-CoV-2 viral load (log value) by the duration of symptom onset of ≤ 3 days on Days 3, 7, and 15- mITT1 population (N=24)

| | | Duration of symptom onset of $\leq 3$ days | | | | | | | |
| --- | --- | --- | --- | --- | --- | --- | --- | --- | --- |
|  |  | Baseline |  | Day 3 |  | Day 7 |  | Day 15/ (EOT) |  |
| Viral Load | Statistics, n(%) | RP7214 (N=11) | Placebo (N=13) | RP7214 (N=11) | Placebo (N=13) | RP7214 (N=11) | Placebo (N=13) | RP7214 (N=11) | Placebo (N=13) |
| Actual | N | 11 | 13 | 8 | 12 | 11 | 13 | 11 | 12 |
| | Mean ± SD | 5.52 ± .19 | 4.51 ± 1.38 | 3.18 ± 1.29 | 3.78 ± 1.39 | 2.54 ± 0.00 | 2.96 ± 1.18 | 3.09 ± 1.33 | 2.98 ± 1.03 |
|  | Median | 5.56 | 5.14 | 2.54 | 3.61 | 2.54 | 2.54 | 2.54 | 2.54 |
|  | Range | (3.8:6.7) | (2.6:6.7) | (2.5:6.1) | (2.5:6.7) | (2.5:2.5) | (2.5:6.7) | (2.5:6.7) | (2.5:5.2) |
| Change from baseline | N | - | - | 8 | 12 | 11 | 13 | 11 | 12 |
| | Mean ± SD | - | - | -2.74 ± 1.57 | -0.65 ± 1.78 | -2.98 ± 1.19 | -1.55 ± 1.74 | -2.43 ± 1.60 | -1.35 ± 1.74 |
|  | Median | - | - | -3.09 | -0.35 | -3.02 | -1.55 | -2.16 | -1.24 |
|  | Range | - | - | (-4.2:-0.5) | (-2.9:3.5) | (-4.2:-1.3) | (-4.2:1.4) | (-4.2:0.0) | (-3.4:2.6) |
| Difference between RP7214 over placebo | Mean ± SD | - | - | -2.09 ± -0.21 | - | -1.43 ± -0.55 | - | -1.08 ± -0.14 | - |
|  | p-value |  |  | 0.0136 | - | 0.0265 | - | 0.1350 | - |

p-value for the viral load was calculated using a two-sample t-test(t).
The reason for the mismatch of N with a total number of 24 patients is due to out-of-window sample collection, data missing, or withdrawn subjects.

Based on vaccination status, RP7214 treated vaccinated patients of the mITT1 sub-population who had symptom onset of ≤ 3 days did not show a statistically significant effect on viral load reduction as compared to placebo. However, RP7214 treated non-vaccinated patients showed statistically significant reduced viral load by a mean of an additional -3.58 (p= 0.0077) log genomic equivalents on Day 3. The reductions on Days 7, and 15 were not statistically significant (**Table 4, Figure 4**).

**Table 4:** Change from baseline in SARS-COV2 viral load (log value) by the duration of symptom onset ≤ 3 days at day 3, 7, and 15 based on vaccination status- mITT1 population (N=24)

| | | Duration of symptom onset of $\leq 3$ days | | | | | | | |
| --- | --- | --- | --- | --- | --- | --- | --- | --- | --- |
|  |  | Baseline |  | Day 3 |  | Day 7 |  | Day 15/(EOT) |  |
| Viral Load | Statistics, n (%) | RP7214 (N=11) | Placebo (N=13) | RP7214 (N=11) | Placebo (N=13) | RP7214 (N=11) | Placebo (N=13) | RP7214 (N=11) | Placebo (N=13) |
| **Vaccinated Patients** |  |  |  |  |  |  |  |  |  |
| Actual | N | 4 | 6 | 4 | 6 | 4 | 6 | 4 | 5 |
| | Mean ± SD | 5.67 ± 1.34 | 4.67 ± 1.56 | 3.82 ± 1.68 | 3.43 ± 0.74 | 2.54 ± 0.00 | 2.54 ± 0.00 | 2.54 ± 0.00 | 2.54 ± 0.00 |
|  | Median | 6.09 | 4.65 | 3.34 | 3.61 | 2.54 | 2.54 | 2.54 | 2.54 |
|  | Range | (3.8:6.7) | (2.6:6.7) | (2.5:6.1) | (2.5:4.3) | (2.5:2.5) | (2.5:2.5) | (2.5:2.5) | (2.5:2.5) |
| Change from baseline | N | - | - | 4 | 6 | 4 | 6 | 4 | 5 |
| | Mean ± SD | - | - | -1.85 ± 1.59 | -1.24 ± 1.36 | -3.13 ± 1.34 | -2.13 ± 1.56 | -3.13 ± 1.34 | -1.72 ± 1.34 |
|  | Median | - | - | -1.35 | -0.89 | -3.55 | -2.11 | -3.55 | -1.55 |
|  | Range | - | - | (-4.2:-0.5) | (-2.9:-0.0) | (-4.2:-1.3) | (-4.2:-0.0) | (-4.2:-1.3) | (-3.4:-0.0) |
| Difference between RP7214 over placebo | Mean ± SD | - | - | -0.61 ± 0.23 | - | -1.00 ± -0.22 | - | -1.41 ± 0.00 | - |
|  | P value | - | - | 0.5518 |  | 0.3112 |  | 0.1632 |  |
| **Non vaccinated Patients** |  |  |  |  |  |  |  |  |  |
| Actual | N | 7 | 7 | 4 | 6 | 7 | 7 | 7 | 7 |
| | Mean ± SD | 5.44 ± 1.20 | 4.38 ± 1.32 | 2.54 ± 0.00 | 4.14 ± 1.85 | 2.54 ± 0.00 | 3.33 ± 1.57 | 3.41 ± 1.62 | 3.30 ± 1.29 |
|  | Median | 4.70 | 5.14 | 2.54 | 3.68 | 2.54 | 2.54 | 2.54 | 2.54 |
|  | Range | (4.1:6.7) | (2.7:5.7) | (2.5:2.5) | (2.5:6.7) | (2.5:2.5) | (2.5:6.7) | (2.5:6.7) | (2.5:5.2) |
| Change from baseline | N | - | - | 4 | 6 | 7 | 7 | 7 | 7 |
| | Mean ± SD | - | - | -3.63 ± 1.07 | -0.05 ± 2.07 | -2.90 ± 1.20 | -1.05 ± 1.85 | -2.03 ± 1.69 | -1.08 ± 2.03 |
|  | Median | - | - | -4.16 | -0.35 | -2.16 | -0.59 | -2.02 | -0.66 |
|  | Range | - | - | (-4.2:-2.0) | (-2.8:3.5) | (-4.2:-1.6) | (-3.2:1.4) | (-4.2:0.0) | (-3.2:2.6) |
| Difference between RP7214 over placebo | Mean ± SD | - | - | -3.58 ± -1.00 | - | -1.85 ± -0.65 | - | -0.95 ± -0.34 | - |
|  | P value | - | - | 0.0077 |  | 0.0503 |  | 0.3626 |  |

p-value for the viral load was calculated using a two-sample t-test(t).
The reason for the mismatch of N with a total number of 24 patients is due to out-of-window sample collection, data missing or withdrawn patients.

**Figure 4:**
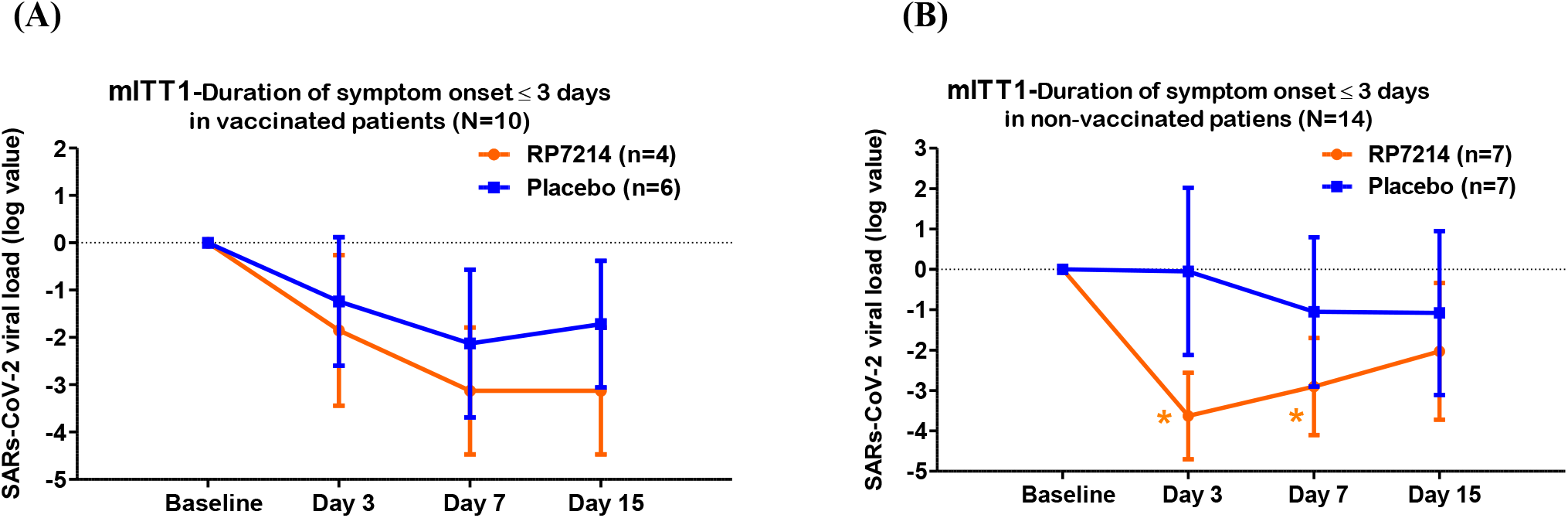
Change from baseline in SARS-CoV-2 viral titer for mITT1 vaccinated patients with duration of symptom ≤ 3 days (A) and mITT1 non-vaccinated patients with duration of symptom ≤3 days (B) Data are presented as means ±SDs. The change in SARS-CoV-2 viral load from baseline to Days 3, 7, and 15 was analyzed using the repeated measure ANCOVA Model. * p-value compares LS means of change from baseline between active and placebo arms.

In mITT1, vaccinated, and non-vaccinated populations having symptom onset of > 3 days, there was no statistically significant difference between the two treatment groups.

There was a trend towards better viral load reduction in RP7214-treated patients with a baseline viral load of 5 log units or higher (mITT2 subpopulation) though it did not achieve statistical significance.

### SARS-CoV-2 viral load clearance (negativity)

The difference in viral load clearance (negativity) was not statistically significant between the RP7214 and placebo groups on Days 3, 7, and 15 in any of the populations. Similarly, time to SARS-CoV-2 viral load clearance (negativity) was not statistically significant between the two treatment groups. When the viral load clearance (negativity) was assessed based on the duration of symptom onset (≤ 3 or > 3 days), the difference in viral load clearance of RP7214 over placebo was not statistically significant for any of the subset populations.

### Symptom resolution

Time to symptom resolution and the proportion of patients with symptom resolution was similar between RP7214 and placebo for all populations.

### Symptom improvement

Time to symptom improvement was similar between RP7214 and placebo groups for all populations. Similarly, the proportion of patients with symptom improvement was similar between RP7214 and placebo for all populations except on Day 3 of the mITT population. The mean of total symptom scores between the two treatment groups across all days was similar. The correlation between change from baseline in viral load and total symptom score was evaluated, however there was no association found between viral load reduction and change in total symptom score.

### Biomarkers

There were no meaningful changes in any of the biomarker levels post-treatment with RP7214 as compared to the placebo in any of the populations.

### Safety

A total of 19 AEs were reported in 17 patients (10.4%), 10 AEs were reported in the RP7214 group, and 9 AEs in the placebo group. Except for one event of anemia, all other AEs were mild in severity The event of anemia was reported in the placebo group and was severe in intensity.

Except for one event of vomiting, all other AEs were considered not related to either RP7214 or placebo. All AEs recovered without sequelae. No drug interruption or discontinuation was reported in any of the patients. No SAEs or deaths were reported during the study (**Tables 5** and **6**). There were no clinically significant changes observed in haematology, clinical chemistry and urine parameters.

**Table 5:** Summary of Overall Adverse Events.

| SOC/PT, n (%) | Study population (N=163) |  |  |
| --- | --- | --- | --- |
|  | RP7214<br>(N=82) | Placebo<br>(N=81) | Total<br>(N=163) |
| Any AE | 10(12.2%) [10] | 7(8.6%) [9] | 17(10.4%) [19] |
| Severity of AEs |  |  |  |
| Mild | 10(12.2%) [10] | 6(7.4%) [8] | 16 (9.8%) [18] |
| Severe | 0(0.0%) [0] | 1(1.2%) [1] | 1(0.6%) [1] |
| The outcome of the event |  |  |  |
| Recovered Without Sequelae | 10(12.2%) [10] | 7(8.6%) [9] | 17(10.4%) [19] |
| Relationship to RP7214 |  |  |  |
| Related | 1(1.2%) [1] | 0(0.0%) [0] | 1(0.6%) [1] |
| Not Related | 9(11.0%) [9] | 7(8.6%) [9] | 16(9.8%) [18] |
| Treatment given for AEs | 5(6.1%) [5] | 5(6.2%) [6] | 10(6.1%) [11] |

Adverse events are represented as patient count (Percentage of patients) [Event Count]

**Table 6:** Summary of Adverse Events by SOC and PT and severity.

| SOC/PT, n (%) | Study population (N=163) |  |  |  |  |
| --- | --- | --- | --- | --- | --- |
|  | RP7214<br>(N=82) |  | Placebo<br>(N=81) |  | Total<br>(N=163) |
|  | Grade 1/2 | Grade 3 | Grade 1/2 | Grade 3 |  |
| Any AE | 10(12.2%) [10] | - | 7(8.6%) [9] | - | 17(10.4%) [19] |
| Blood and lymphatic system disorder | 1(1.2%) [1] | - | 2(2.5%) [2] | - | 3(1.8%) [3] |
| • Anemia | 1(1.2%) [1] | - | - | 1(1.2%) [1] | 2 (1.2%) [2] |
| • Neutrophilia | - | - | 1(1.2%) [1] | - | 1(0.6%) [1] |
| Gastrointestinal disorders | 7(8.5%) [7] | - | 5(6.2%) [5] | - | 12(7.4%) [12] |
| • Abdominal pain upper | 1(1.2%) [1] | - | - | - | 1(0.6%) [1] |
| • Diarrhoea | 1(1.2%) [1] | - | 2(2.5%) [2] | - | 3(1.8%) [3] |
| • Nausea | 1(1.2%) [1] | - | 2(2.5%) [2] | - | 3(1.8%) [3] |
| • Vomiting | 4(4.9%) [4] | - | 1(1.2%) [1] | - | 5(3.1%) [5] |
| Nervous system disorders | 2(2.4%) [2] | - | 1(1.2%) [1] | - | 3(1.8%) [3] |
| • Headache | 2(2.4%) [2] | - | 1(1.2%) [1] | - | 3(1.8%) [3] |
| Respiratory, thoracic, and mediastinal disorders | - | - | 1(1.2%) [1] | - | 1(0.6%) [1] |
| • Oropharyngeal pain | - | - | 1(1.2%) [1] | - | 1(0.6%) [1] |

Adverse events are represented as Patient count (Percentage of patients) [Event Count]

## Discussion

SARS-CoV-2infection led to a worldwide pandemic and is of immense global public health concern.^[1]^ Although the development of vaccines against SARS-Cov-2 has been able to create a significant impact on reducing the risk of hospitalization and death due to Covid, there is still an emerging need for therapeutics. There are still populations in the world that have not vaccinated themselves due to cultural, co-morbidities, age, and access-related issues. These populations are especially vulnerable and hence a therapeutics-based approach is crucial. Even in populations that have been vaccinated, an infection can still occur although not many instances are severe and require hospitalization. In a vulnerable vaccinated population which gets infected, there is a risk of disease progression, and a therapeutic intervention will be required to stop disease progression especially in patients with co-morbidities. ^[15, 16]^

While inhibitors of DHODH have shown efficacy in malaria and autoimmune diseases such as rheumatoid arthritis and psoriasis, human trials are ongoing to elucidate their effect on viral replication in patients with Covid-19 infection.^[17]^

In this study, we planned to enroll 204 patients, but could only enroll 163 patients due to the waning wave of the pandemic. Based on the time period during which the study started to when enrolment was closed, the pandemic in India was largely dominated by the Omicron variant of SARS-Cov-2 (*though the variant was not genomically tested in the study)*.^[18]^ Our data largely reflects the effect of RP7214 in an Omicron infected population which has a rapid course of infectivity and decline as well with a very low risk of converting to severe disease requiring hospitalization.^[18]^ The data from this study was thus largely in a population who were at low risk of severe disease. Given this data, our results did not show any severe disease or requirement of hospitalization in either of the treatment arms.

RP7214 was given at a dose of 400 mg BID. This dose was selected based on the PK seen in the phase I study. Since we specifically enrolled patients with at least one co-morbidity, the viral kinetics in these patients were presumed to be different from the patients without co-morbidities.^[19, 20]^ Hence a treatment duration of 14 days with the highest dose of 400 mg BID tested in the phase I study was used with the intent to expose patients to RP7214 for an additional 7-8 days post achievement of steady-state level to optimise the response to the treatment. Other antiviral agents for SAR-CoV-2 infection such as remdesivir and favipiravir have treatment courses ranging from 10 to 14 days duration. ^[21, 22]^ Hence a treatment duration of 14 days was considered, taking into account the duration of the disease and potential virus shedding that occurs during the course of the disease.

RP7214 did not show a statistically significant difference in viral load reduction compared to the placebo in the planned analysis population dataset. However, there was a statistically significant reduction in viral load in the subset of patients who started treatment at an early stage of the disease (symptoms onset ≤ 3 days). This dataset was further analysed for patients who received prior vaccination. Data showed that in patients who had not received prior vaccination and were treated with RP7214 early on (≤ 3 days) showed maximal viral load reduction. In the population showing a significant difference in viral load as compared to placebo, there was no concurrent reduction in symptom resolution time or any change in biomarkers as compared to placebo. This could be due to the pathophysiology of the Omicron variant where though the level of infection could be high, the resolution of clinical symptoms and associated markers is rapid. This could be the reason for no difference being seen in clinical symptoms and biomarkers across the treatment groups.

Patients who have had prior vaccinations would still have persistent antibodies in circulation,^[23]^ though we could not record the date of all vaccinated patients. Since the unvaccinated patient population was a more vulnerable group and with the majority of them having a *de novo* infection, RP7214 was able to show an impact in reducing the viral load and leading to a faster reduction in viral elimination kinetics as compared to placebo. Also in this population, which had an early start of treatment, RP7214 was able to show an effect when the viral load peaked and hence there was a steeper fall as compared to the placebo.

Based on the anti-inflammatory effects of RP7214 in the lung seen in pre-clinical models, (data in the file) we expected that there would be an improvement in the time of resolution of Covid 19 respiratory symptoms. However, there was no difference from the placebo. The lower virulence of the Omicron variant,^[18]^ could be the reason for not achieving a clinical symptom-based benefit. We believe this could have been carved out better in a prior variant like the delta variant.

Additionally, there was a trend towards better viral load reduction in RP7214-treated patients with a baseline viral load of 5 log units or higher as compared to placebo though not statistically significant. This may show some potential of RP7214 in patients who have been exposed to high viral loads especially in areas which are quarantined or in healthcare setups.

RP7214 was well tolerated and there were no safety concerns during treatment for as long as 14 days. Based on the study results, we believe that there is an opportunity for further development of RP7214 in SARS-CoV2 in vulnerable populations and will prevent the risk of severe disease in this group of patients.

Limitations of the study were a lesser virulent variant of Omicron, a single-dose regimen and not being able to recruit the entire set of patients as envisaged in the study due to the waning of the pandemic.

## Conclusions

RP7214 at 400 mg BID dose level showed a statistically significant reduction in viral load at an early stage of the disease (symptoms onset ≤ 3 days) and in patients who were not vaccinated. Though not statistically significant, there was a trend towards better viral load reduction in RP7214-treated patients with a baseline viral load of 5 log units or higher. Overall, RP7214 at 400 mg BID dose level was safe and well tolerated and showed a favorable safety profile.

## Data Availability

The data that support the findings of this study are available from the corresponding author Ajit Nair upon reasonable request.

## Acknowledgements

We are grateful to all the patients who participated in this study and their families. We would like to thank the site research staff, site scientific staff, central laboratory, and data safety monitoring board. We thank JSS Medical Research Asia Pacific Private Limited, Faridabad, India and the study teams for monitoring and management.

## Notes

**Declaration of Conflicting Interests** Prajak Barde, Kasi V Routhu, and Ajit Nair are employed at Rhizen Pharmaceuticals. Swaroop V. Vakkalanka has equity, ownership & employment at Incozen therapeutics Pvt Ltd. All authors retained full ownership of the manuscript content and approved the final draft for submission

**Data Availability Statement** The data that support the findings of this study are available from the corresponding author Ajit Nair upon reasonable request.

**Funding** This work was supported by Incozen Therapeutics Pvt Ltd., Lab Suites 231-234, Building 450, MN Science and Technology Park, Genome Valley, Turkapally (V), Shameerpet Mandal, Hyderabad-500078, Telangana, India

### Competing Interest Statement

The authors have declared no competing interest.

### Clinical Trial

NCT05007236

### Funding Statement

This work was supported by Incozen Therapeutics Pvt Ltd., Lab Suites 231-234, Building 450, MN Science and Technology Park, Genome Valley, Turkapally (V), Shameerpet Mandal, Hyderabad-500078, Telangana, India

### Author Declarations

The Institutional Human Ethics Committee of Panimalar Medical college hospital and research Institute, Chennai, India gave the first ethics approval for this work

